# Deep learning-based prediction of one-year mortality in the entire Finnish population is an accurate but unfair digital marker of aging

**DOI:** 10.1101/2023.09.18.23295726

**Authors:** Andrius Vabalas, Tuomo Hartonen, Pekka Vartiainen, Sakari Jukarainen, Essi Viippola, Rodosthenis S. Rodosthenous, Aoxing Liu, Sara Hägg, Markus Perola, Andrea Ganna

## Abstract

**Background:** Accurately predicting short-term mortality is important for optimizing healthcare resource allocation, developing risk-reducing interventions, and improving end-of-life care. Moreover, short-term mortality risk reflects individual frailty and can serve as digital aging marker. Previous studies have focused on specific, high-risk populations. Predicting all-cause mortality in an unselected population incorporating both health and socioeconomic factors has direct public health relevance but requires careful fairness considerations.

**Methods:** We developed a deep learning model to predict 1-year mortality using nationwide longitudinal data from the Finnish population (*N* = 5.4 million), including >8,000 features and spanning back up to 50 years. We used the area under the receiver operating characteristic curve (AUC) as a primary metric to assess model performance and fairness.

**Findings:** The model achieved an AUC of 0.944 with strong calibration, outperforming a baseline model that only included age and sex (AUC = 0.897). The model generalized well to different causes of death (AUC > 0.800 for 45 out of 50 causes), including COVID-19 which was not present in the training data. The model performed best among young females and worst in older males (AUC = 0.910 vs. AUC = 0.718). Extensive fairness analyses revealed that individuals belonging to multiple disadvantaged groups had the worst model performance, not explained by age and sex differences, reduced healthcare contact, or smaller training set sizes within these groups.

**Conclusion:** A deep learning model based on nationwide longitudinal multi-modal data accurately identified short-term mortality risk holding the potential for developing a population-wide in-silico aging marker. Unfairness in model predictions represents a major challenge to the equitable integration of these approaches in public health interventions.

## INTRODUCTION

Explaining variability in individuals’ aging trajectory, life expectancy and mortality risk remains a fundamental task for public health, medical research, and policymaking^1,2^. Adequate identification of individuals at risk of short-term death is fundamental for planning risk-reducing interventions. Short-term mortality prediction is of particular value to improve the quality of care at the end of life, and at the same time reduce costs via optimization of healthcare resource utilization^3^. It is also important to understand the progressive mismatch that arises between an individual’s chronological age and biological age, particularly at the most advanced ages as this mismatch has a large effect on our capacity to predict mortality^2,4,5^. Overall, understanding the sources of increased biological heterogeneity in old age remains a central question in aging research^4^.

Recent advances in machine learning, coupled with the wider availability of digitized medical and socioeconomic information at a population level, have paved the way for the development of algorithms that can predict patients’ future health trajectories and aid medical decision-making^6,7^. Deep learning (DL) models can leverage massive amounts of data, requiring minimal pre-processing or feature engineering. A clear advantage of DL models is the possibility to analyse an individual’s longitudinal history, considering time intervals elapsed between different events, including medical encounters as well as socioeconomic information.

Unlike traditional statistical methods, DL is often viewed as a “black box” meaning that its decisions are difficult to interpret. While existing explainability methods can provide insights into which attributes are important at a level of an individual, they do not facilitate the understanding of differences in predictions across groups of individuals^8^. Understanding how model performance varies across different groups becomes especially important when considering issues of fairness. Fair algorithms should not exhibit bias or preference towards any individual or group based on inherent or acquired attributes^9^. There have been instances where DL algorithms are unfair^10^, particularly when they perform poorly for socially disadvantaged individuals, who may face higher barriers to accessing healthcare, resulting in more missing data and measurement errors that ultimately skew the predictions^11^.

For instance, Fong et al.^12^ found that a model predicting hospital readmissions achieved much higher prediction accuracy among self-reported Caucasian individuals compared to other racial and ethnic groups. Similarly, Meng et al.^13^ identified disparities in the frequency of mechanical ventilation interventions across different ethnicities, sexes, and ages leading to differences in prediction accuracy across groups. Chen et al.^14^ found that prediction models performed worse for males compared to females and among individuals with public, rather than private, health insurance.

Our study aims to accurately predict one-year mortality for every Finnish resident by utilizing comprehensive, nation-wide multi-modal information and to evaluate how the prediction accuracy varies within different groups defined by health, geographical location, and socioeconomic characteristics. To achieve this objective, we developed a state-of-the-art DL model. Because (bio)markers of aging are suggested to predict future health and survival better than chronological age we compared its performance with that of a simpler baseline model, which considers only age and sex. While previous studies have attempted to predict short-term mortality using electronic health records^15^, environmental and lifestyle factors^16^, and biomarkers^17^, our study significantly contributes to the field in several aspects.

Firstly, our study includes the entire Finnish population, resulting in a large and population-representative sample. Secondly, our study utilizes an unprecedented number of longitudinal data modalities with comprehensive and high-quality data collected in national registers. Particularly noteworthy is extensive socioeconomic information which was limited in previous studies. Finally, with these two advantages, we were able to explore differentiable predictions and fairness at a level of detail not previously possible, for example by leveraging detailed economic measurements to identify disadvantaged individuals, thus contributing to the understanding of disparities in healthcare predictions.

## RESULTS

### Individuals included in the study, data, and model

FinRegistry (https://www.finregistry.fi/) is a comprehensive register-based data resource that provides access to a diverse range of health and sociodemographic data for the entire Finnish population. The unique characteristic of this resource is the breadth of data modalities included: healthcare visits, health conditions, medications, surgical procedures, demographic characteristics, welfare benefits, pensions and detailed socioeconomic information (**Supplementary materials** provide a detailed description of data sources).

Notably, some of this information spans decades, dating as far back as the 1970s. The Causes of Death registry is particularly relevant to this study as it offers comprehensive information about death events and causes of death.

FinRegistry covers all Finnish residents on January 1, 2010, as well as their parents, spouses, children, and siblings. For our study, we included all individuals alive and not emigrated on January 1, 2020 (*N* = 5,364,032, **Figure 1A**, for a detailed study overview). Our objective was to predict all-cause mortality within one year, with approximately 1% of individuals dying within this timeframe. To ensure the generalizability of our predictions, we considered three consecutive years for training, validation, and testing. Specifically, we predicted mortality in 2018 during training, in 2019 for validation, and in 2020 for testing.

**Figure 1:**
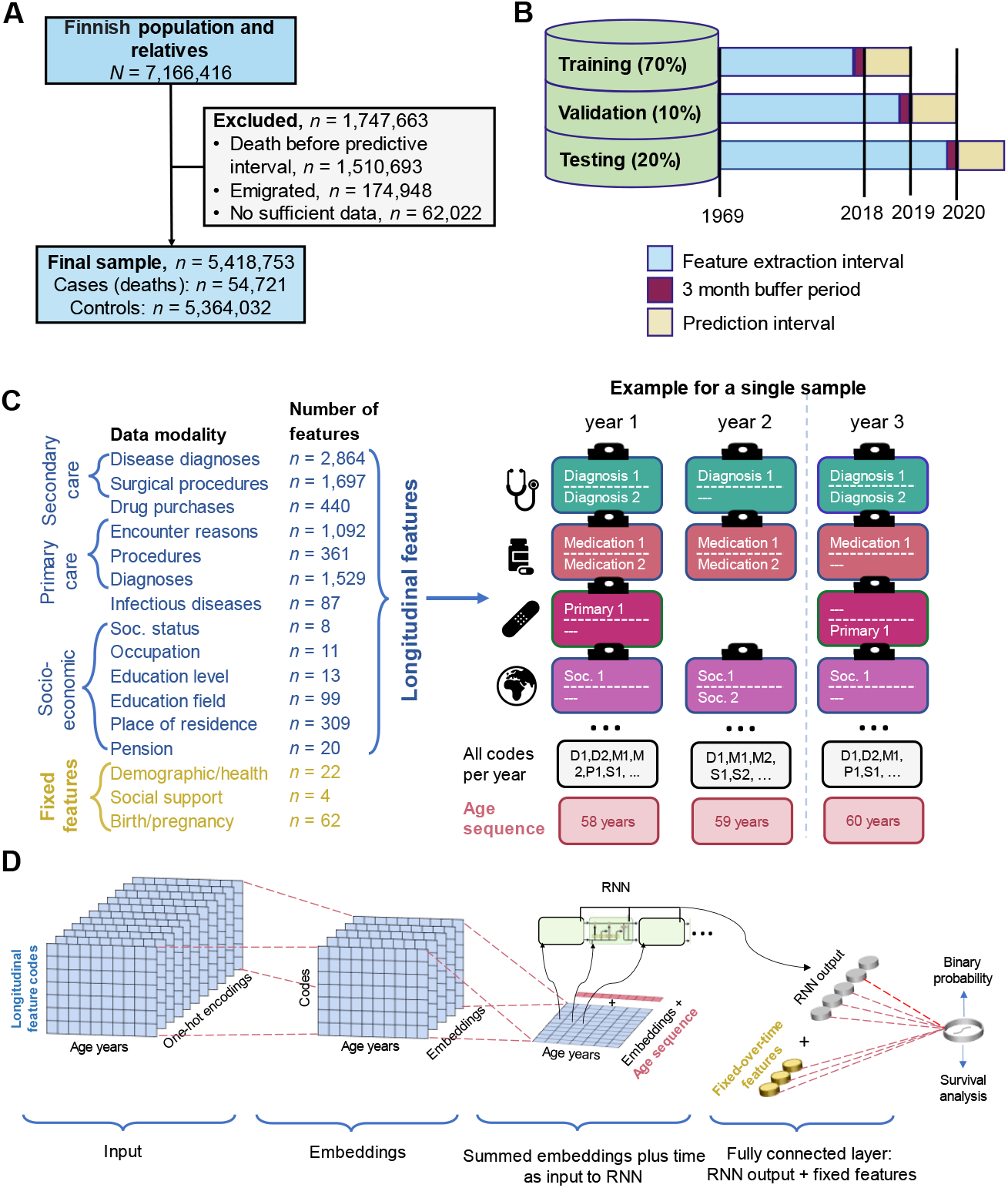
Study population, data, and model. (A) Study population and inclusion/exclusion criteria. (B) Data division into the training, validation and testing sets in a prospective fashion. (C) Features included in the model, either treated longitudinally or fixed over time (different types of features/model inputs are colour coded in panels C and D) with an example of longitudinal features available for an individual across three years. *n* denotes the number of features in different modalities. (D) A graphical representation of the RNN model. Longitudinal records were embedded and then together with an age sequence used as inputs for a recurrent layer. Fixed over time features were also added before the output layer. Soc. = Socioeconomic.

These shifts ensured that the validation and testing prediction periods remained “unseen” to the model during training (**Figure 1B**). The COVID-19 pandemic has significantly disrupted the healthcare system in 2020. Therefore, using this year for predictions in our model serves as a rigorous “stress test” for assessing its robustness.

To build our models, we employed both fixed over time and longitudinal features (**Figure 1C**). Longitudinal features used coded records exactly as they appeared in the registers while preserving temporal information on the duration between different events. Fixed over time features were only used to capture information that remained constant throughout an individual’s lifetime, such as basic demographic information. By combining both types of features, we were able to capture both the dynamic and static characteristics of each individual, improving the predictions. Overall, we included 8,620 features, of which 90 were fixed over time and 8,530 were longitudinal.

To capture the complex interactions between events over time, we utilized a recurrent neural network (RNN) with a gated recurrent unit (**Figure 1D**). RNNs have been proven effective in modelling patients’ health histories^18^ and have demonstrated comparable performance to other sequential deep learning models, such as transformers, in predicting clinical events^19,20^.

To evaluate our deep learning model against a simpler baseline model, we employed a logistic regression model that included only age and sex as predictors of mortality.

### Descriptive results

We explored age and sex distribution in our data as crucial factors influencing mortality (**Figure 2A, B**). The mean age of our study population was 44.4 years on January 1, 2020, and there were more females (50.8%) than males (49.2%). The mean age at death, in 2020, was 79.7 (83.3 for females and 76.1 for males) and only 13.2% of deaths occurred before 65 years of age.

**Figure 2:**
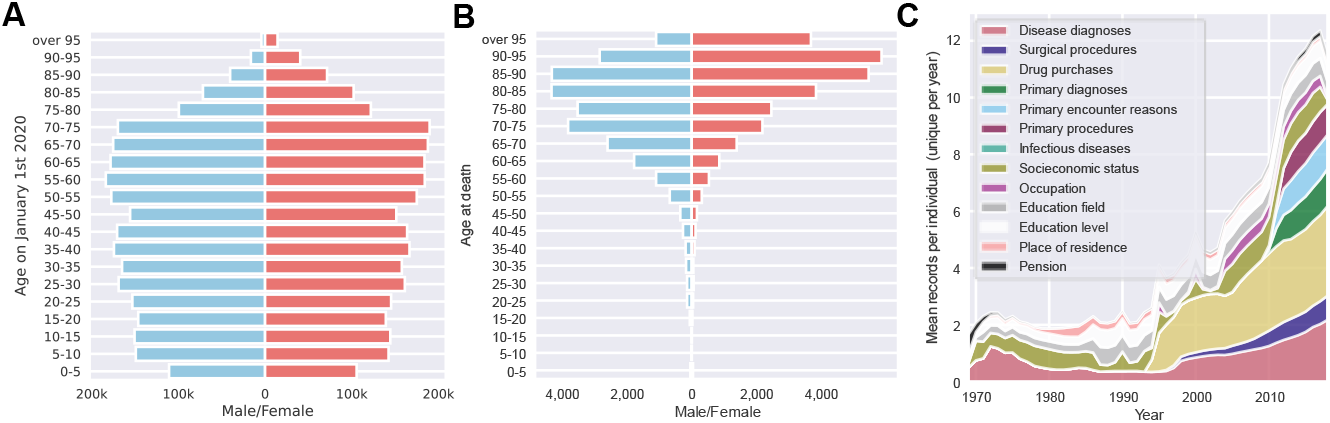
Descriptive results in the testing set. (A) A pyramid plot showing age and sex distribution for a full study population at the start of the predictive interval on January 1, 2020. (B) A pyramid plot showing age and sex distribution for individuals who did die during the predictive interval (the year 2020). (C) Distribution of the average yearly number of records per individual over time within the testing set. For each individual duplicate records within a single year were not included.

We explored the amount of longitudinal data available over time (**Figure 2C**). There was a gradual increase in the mean number of records available per individual over time, with some data modalities starting in later years. Specifically, the drug purchase register was introduced in 1995, followed by the outpatient register (reflected in disease diagnoses and surgical procedures categories) in 1998, and finally the primary care register in 2011.

Overall, most individuals had information from multiple modalities, with 78% of individuals having records for at least 8 modalities (**Supplementary Figure 1**).

### Model performance

The RNN model included 2.9 million trainable parameters and achieved an area under the receiver operating characteristic (AUC) of 0.944 (95% confidence intervals (CI) 0.942 to 0.946) for binary classification, surpassing the baseline model that relied solely on age and sex, which achieved an AUC of 0.897 (95% CI 0.894 to 0.899, **Figure 3A**). Additionally, the RNN model exhibited superior calibration, as indicated by a lower mean squared error between predicted values and true labels (**Figure 3B**). The RNN model achieved a higher area under the precision-recall curve (AUPRC) than the baseline model (0.223 vs. 0.119, **Figure 3C**). It’s worth noting that AUPRC is influenced by the degree of class imbalance and is expected to be lower in situations where the class imbalance is high, as observed in our study.

**Figure 3:**
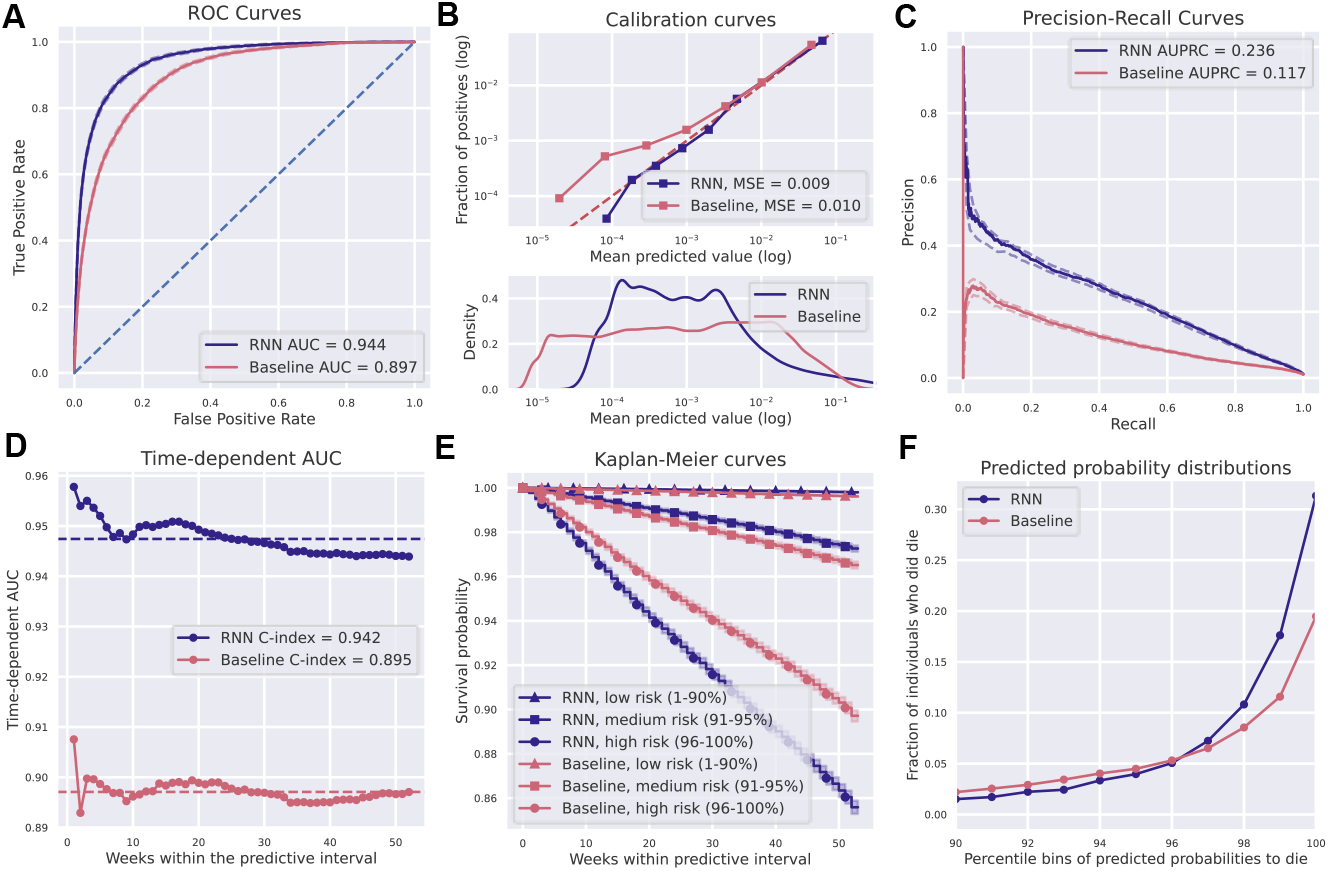
One-year mortality prediction results. (A) AUC for the recurrent neural network (RNN) and baseline models. (B) Calibration curves for RNN and baseline models. Observed and predicted probabilities of death for each risk decile are reported on a log scale because of a skewed probability distribution, with most values close to zero. This is evident in the bottom panel showing predicted probability densities for both models. A quantile binning strategy for calibration curves was used to assure an equal number of samples in each bin. (C) Precision-recall curves and AUPRC values for the two models. (D) Time-dependent AUC curves and C-indexes for each of the 52 weeks in the predictive interval. Dashed lines show the mean AUC for each model. (E) Kaplan-Meier curves for predicted low, medium, and high mortality risk groups in the testing set for two models. Stratification of individuals to the risk groups is according to their predicted survival over time within the predictive interval. Although the low-risk group covers a large 1-90 percentile range, the curves are nearly horizontal and overlap, with low mortality over time for both models. (F) Fractions of individuals who did die in the testing set as a function of percentile bins of predicted mortality probabilities within the predictive interval for the two models. We only plot individuals at medium and high risk (90+ percentile).

When we considered time-to-death rather than binary classification, the RNN yielded a C-index of 0.942 (95% CI 0.940 to 0.944). While the RNN model demonstrated slightly better performance in predicting mortality at the start of the year, it maintained a consistently high C-index throughout the entire predictive interval (**Figure 3D**).

We compared the Kaplan-Meier (KM) curves for three risk groups categorized by the predicted mortality probability from either the RNN or the baseline model (**Figure 3E**). The RNN model showed a larger disparity in survival rates among the three groups, compared to the baseline model. For instance, the high-risk group, comprising individuals with predicted mortality probabilities ranging from the 96th to 100th percentile (i.e., 5% of the individuals with the highest predicted risk), exhibited a mortality rate of 16.8% by week 52, compared to 11.4% predicted by the baseline model (**Figure 3F**). To put it differently, the RNN model predicted 69.5% of all deaths that occurred in the testing set to be in the high-risk group, compared to the baseline model’s prediction of 49.6% of all deaths Overall, the RNN model outperformed the baseline model in differentiating between medium and high-risk groups.

When evaluating the importance of different modalities for mortality prediction, longitudinal features had greater predictive power than fixed over time features (**Supplementary Figure 2**). Among longitudinal features, medical data were more important than socio-demographic data (AUC 0.942 vs. 0.922). The two top-performing modalities, disease diagnoses and drug purchases achieved an AUC of 0.936 and 0.935, respectively.

### Model performance by cause of death and age

To test the robustness of the model across different medically and socioeconomically relevant groups we first examined groups based on different causes of death and age. We took two different approaches.

The first approach is **group identification**, which evaluates the predictability or identifiability of a specific subgroup *within the entire population*. Previous studies have used this approach to compare the predictability of different diseases^21^, or the subtypes of diseases^22,23^ within the pool of healthy individuals.

The second approach is **group differentiation**, which compares prediction performance *within a particular subgroup of the population* relative to another subgroup from the same population (e.g., a specific age group). This approach is typically used in algorithm fairness studies to assess differences in prediction performance between groups defined by ethnicities, sexes, ages, and other attributes. Aging researchers also utilize this approach to evaluate the efficacy of biological age predictors beyond what is solely accounted for by chronological age in different age groups^15,24^.

We employed the group identification approach to compare mortality prediction across 50 different causes of death (COD, five causes of death were excluded due to an insufficient number of cases of 5 or fewer, **Figure 4A**). The frequency of different CODs varied significantly, ranging from less than 1% for external CODs (such as accidents or suicides) to 15.8% and 18.8% for the most common CODs, namely ischemic heart disease and dementia, respectively (rightmost part of **Figure 4A**). The RNN model showed good predictive performance across CODs, achieving an AUC of over 0.8 for 45 out of 50 CODs. Prediction performances for CODs related to accidents and violence were significantly lower than disease-related CODs (average AUC of 0.761 and 0.939, respectively). Nonetheless, the RNN model substantially outperformed the baseline model, especially for CODs related to accidents and violence with an average AUC improvement of 0.11 (light blue bars in **Figure 4A**). It is worth noting that COVID-19 emerged as a novel cause of death in 2020, and although the RNN model was not designed to predict COVID-19 mortality due to the absence of COVID-19 deaths in the training data, it achieved a high AUC of 0.956.

**Figure 4:**
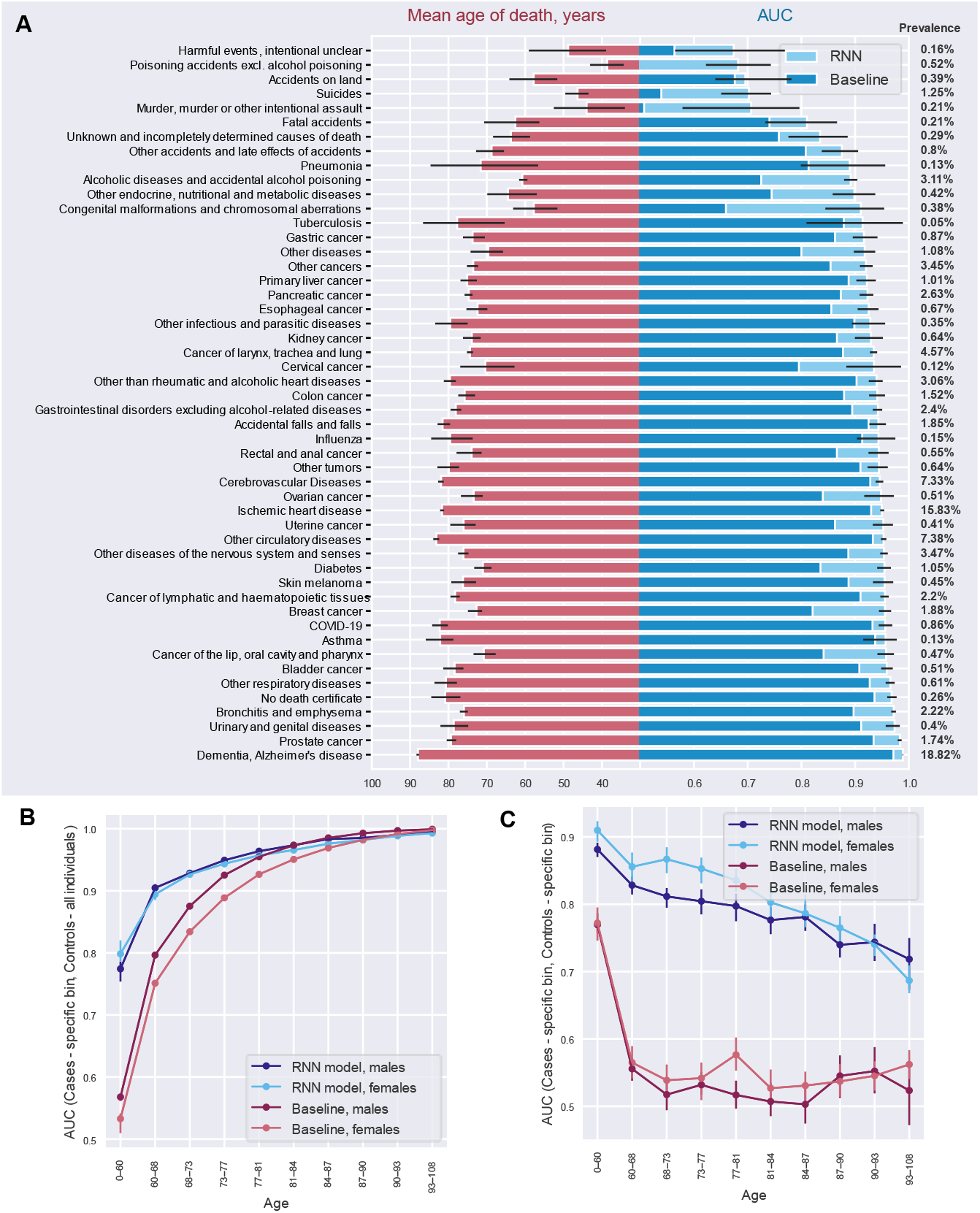
Mortality prediction for the cause of death, age, and sex subgroups. (A) The left side of the panel provides the average age at the time of dying from a specific cause within a testing set. The right side of the panel provides area under the receiver operating characteristic curve (AUC) values for individuals dying from a specific cause. AUCs are reported separately for the recurrent neural network (RNN) and baseline models. Confidence intervals obtained via bootstrapping are reported only for the RNN model to improve readability. The percentage of people dying from a specific cause is given in the right margin of a panel. Only causes of death with 20 or more cases were considered. (B) Model AUC scores for specific age/sex subgroups of cases (group identification approach: considering cases from a specific age/sex subgroup versus all controls). (C) Model AUC within specific age/sex subgroups (group differentiation approach: considering cases and controls from a specific subgroup only). This corresponds to evaluating the predictive performances of the model after the effect of age as a predictor has been substantially minimized (for more precise removal of age effect, refer to **Supplementary Figure 3**). In panels, B-C, ten age bins were used assuring an equal number of cases in each. The error bars indicate 95% confidence intervals computed using bootstrapping.

Both the RNN and baseline models demonstrated better predictions for CODs occurring at older ages. For instance, individuals who died from dementia at a mean age of 87.9 years were well predicted by both models (AUC = 0.989 and 0.971 for RNN and baseline, respectively). Conversely, the RNN model was substantially better at predicting CODs occurring among younger individuals. For example, suicide (mean age of 46.3) was substantially better predicted by the RNN compared to the baseline model (AUC = 0.702 vs. AUC = 0.539). Overall, the mean age at death was the primary factor contributing to differences in AUC for the baseline model (*R*^*2*^ = 0.992), whereas this association was weaker for the RNN model (*R*^*2*^ = 0.809). Interestingly, there was no discernible relationship between the prevalence of each COD and the prediction performance, as both rare and common causes of death achieved high AUCs (*R*^*2*^ = 0.091 and 0.057 for the baseline and RNN models, respectively).

As COD predictability showed a strong correlation with age, we further explicitly explored the relationship between model performance and age at death. We employed both group identification (**Figure 4B**) and group differentiation (**Figure 4C**) approaches to explore this relationship in detail and compare the approaches.

First, using the group identification approach, we explored how well a model was able to identify individuals who died within a specific age bin among the entire population irrespective of their age. The results mirrored those of the COD analyses, with both the RNN and baseline models exhibiting better predictions for older age groups. The RNN model performed notably better, particularly in the youngest individual bins (**Figure 4B**).

Second, we used a group differentiation approach and assessed model performance limiting cases and controls to a specific age bin (**Figure 4C**). This corresponds to evaluating the predictive performances of the model after the effect of age as a predictor has been substantially minimized. In contrast to the group identification task, the RNN model’s prediction performance declined in older age bins, showing higher performance for young females than young males. For the baseline model, performance was at a random guessing level (AUC ∼ 0.50) in each age bin, except for the youngest age group with the widest age range and not sufficient control for age differences between cases and controls. After exactly matching the age and sex of cases and controls within each age group, the baseline model, but not the RNN model, showed random guessing level performance across all age groups (**Supplementary Figure 3**).

### Prediction fairness

We examined the fairness of predictions by comparing model performance across groups of individuals based on geographic location, monthly pension level and other sociodemographic variables.

First, we compared the RNN model performance across different regional municipalities. We found significant variability in prediction performance between different regional municipalities, with AUCs ranging from 0.881 to 0.964 (**Figure 5A**). For example, we observed lower prediction performance in the northern Lapland region, consisting of six regional municipalities, compared to the rest of Finland (AUC = 0.924 vs. 0.939, *p* = 0.002). Substantial differences were observed between neighbouring regional municipalities. For example, Pohjois-Satakunta and Luoteis-Pirkanmaa, despite their geographical closeness, had significantly different model performances (AUC = 0.964 versus 0.890, *p* < 0.001). The differences were partly explained by population density as we observed a positive correlation (*r* = 0.23, *p* = 0.05) between population density and AUC in different regional municipalities. To determine whether the observed variability in AUC was influenced by the model’s awareness of geographic information, we retrained the RNN model without geographic features, but we still observed similar differences in performance (**Supplementary Figure 4**). The baseline model had higher variability in its prediction performance across different regional municipalities compared to the RNN model (standard deviation in AUC of 0.027 vs. 0.016, **Figure 5B**).

**Figure 5:**
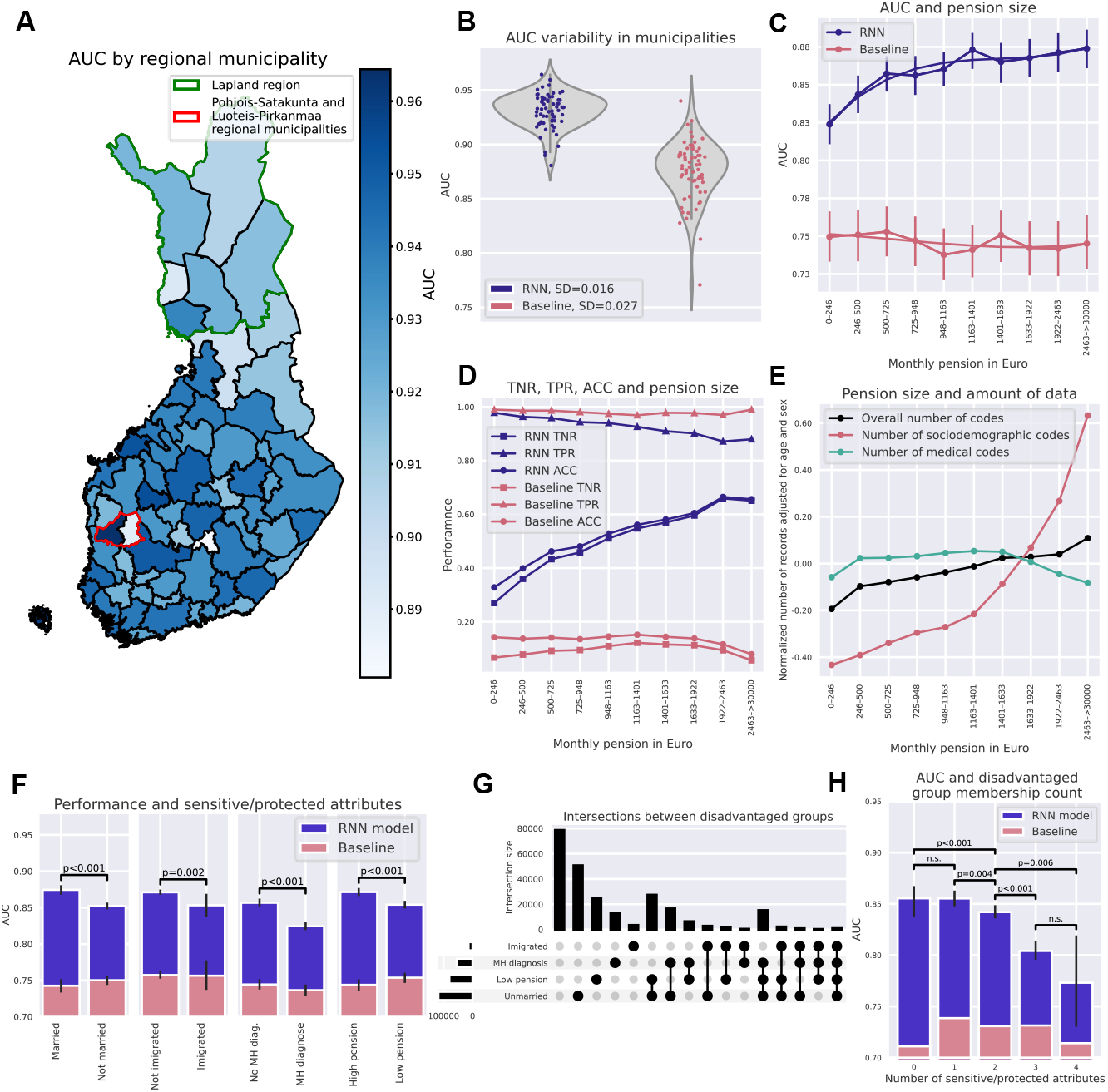
Fairness regarding the place of residence, pension size, and other sensitive attributes. (A) AUC variation by a regional municipality in Finland. The green border marks the Lapland region in which AUC was significantly lower than in the rest of Finland, red border surrounds two neighbouring regional municipalities with significantly different AUCs. Two regional municipalities Mariehamns stad and Ålands skärgård are not plotted because the minority class had fewer than 20 samples. (B) Each dot represents a different regional municipality. Variability in prediction performance in different regional municipalities shows a larger spread and greater geographic variability for baseline compared to the RNN model. (C) AUC from baseline and RNN models within each pension level bin. The RNN model has higher prediction performances among individuals with a higher pension. (D) Accuracy (ACC), sensitivity (TPR) and specificity (TNR) for the RNN and baseline model as a function of pension. The classification metrics were calculated based on a probability cut-off of 0.0089 for the RNN model and 0.0094 for the baseline model (see Methods for cut-off calculation). For an RNN model, an increase in AUC with greater pension size was driven by TNR – better identification of individuals who did not die during a predictive interval. (E) The average number of total records available for training the RNN model as a function of pension size. The average number of total records per individual is adjusted for age and sex and then normalized. This metric allows evaluation of whether individuals with a higher pension have more information available, potentially explaining the better performance of RNN models. Records from three main data modalities are reported. In panels, C-E, ten pension bins were used assuring an equal number of cases in each. (F) AUC for RNN and baseline model for different attributes considered protected or sensitive: marital status, immigration status, mental health (MH) diagnosis, and pension size (individuals were split into two pension size groups assuring an equal number of cases in each). Statistical significance was assessed using permutation testing. (G) UpSet plot^25^ visualizes intersections between four groups of disadvantaged individuals. (H) AUC for RNN and baseline model in individuals having none, one, or several disadvantages across four sensitive/protected attributes simultaneously. Statistical significance was assessed using permutation testing.

Second, we investigated the fairness of our mortality prediction model with respect to average monthly pension levels in 2020. We chose old-age pension as it is based on an individual’s income throughout their working life and is particularly relevant among older individuals, where most deaths occurred. To focus our analysis, we limited our investigation to individuals over 65 years of age, as this group accounted for 85% of all deaths in 2020, and 93% in this group received an old-age pension. There was a clear positive relationship between pension levels and AUC for the RNN model, with higher AUC for higher pension (e.g. AUC = 0.824 for pension between 0 and 246 € month vs. AUC = 0.874 for pension between 2,463 and > 30,000 € month, p<0.001). No such relationship was observed for the baseline model (**Figure 5C**). Similar results were also observed after matching individuals for age and sex within each pension bin (**Supplementary Figure 5**) and when a model was retrained without pension features (**Supplementary Figure 6**). Analysis of sensitivity (TPR) and specificity (TNR) revealed that the increase in AUC with greater pension size was predominately driven by increasing TNR (i.e., better identification of individuals who did not die during the predictive interval, **Figure 5D**). We also explored whether differences in the amount of training data could have influenced AUCs in different pension bins (**Figure 5E**). Individuals in higher pension bins tended to have more socioeconomic records while the number of medical records and an overall number of records remained similar within different pension bins.

Third, we expanded our fairness analyses to four sensitive/protected attributes, which partially overlapped (**Figure 5G**). We found that AUCs for sensitive/protected groups, such as those who were unmarried, had immigrated, had mental health diagnoses, or received low pensions, were significantly lower than for their counterparts (**Figure 5F**; *p* < 0.002 for all comparisons). We also performed the same comparisons after matching for age and sex, within socially disadvantaged and advantaged groups: the observed effects remained, except for immigration status (**Supplementary Figure 7**). Additionally, belonging to multiple sensitive/protected groups simultaneously resulted in considerably worse AUCs (**Figure 5F**) in the RNN, but not in the baseline model.

## DISCUSSION

In this study, we used a nationwide, high-quality multimodal dataset to predict one-year all-cause mortality for the entire Finnish population and to investigate variability in predictions and fairness at a level of detail not previously possible.

Despite using a prospective testing approach to ensure the prediction period remained “unseen” by the model during training, our model exhibited strong predictive abilities (AUC = 0.944 (95% CI 0.942 to 0.946), and was well-calibrated, surpassing a simpler baseline model. For example, a significant proportion of all deaths (69.5%) occurred in a high-risk group comprising only 5% of individuals with the highest predicted risk.

Due to the small number of deaths per year, accurately predicting one-year mortality in the general population necessitates a significant sample size. However, to our knowledge, there have been no previous attempts at nationwide prediction with earlier studies focusing solely on high-risk individual groups, such as elderly patients in care homes^26^. Our model, on the other hand, can be flexibly applied across different ages and cause of death groups, including previously unseen causes of death such as COVID-19. Notably, our model demonstrated a significant improvement over the baseline model when predicting deaths resulting from accidents or violence. We speculate that the inclusion of socioeconomic features may have aided in predicting such seemingly external causes of death.

Our model’s advantages became apparent when evaluating its prediction performance beyond chronological age. Even after removing the age effect, which is the strongest mortality predictor, our model achieved an AUC of 0.769 for males and 0.822 for females aged 0 to 60 (**Supplementary Figure 3**). This additional predictive performance beyond age suggests the potential of our model as a digital marker of biological age, which is distinct from chronological age. In comparison, markers of biological aging, such as frailty indexes, DNA methylation and telomere length, achieve lower performance for mortality prediction^27–29^. Intriguingly, our model exhibited stronger predictive performance among younger, but not older females, compared to males. As we observe greater contact with healthcare among younger females, partially due to childbirth compared to males (**Supplementary Figure 8**), we speculate that this may provide predictive information that is not available for males.

After controlling for the age effect, our model’s performance gradually decreased in older individuals. As people age, they start to differ more from each other because they experience biological and environmental changes at varying rates and degrees^4^. This increases variability in functional abilities, such as mobility, self-care, ability to perform usual activities, pain/discomfort, and anxiety/depression^5^. Furthermore, the combination of increased damage and reduced resilience can lower the threshold for adverse events to result in mortality^2^. The presence of significant heterogeneity among older individuals likely diminishes the distinctiveness of data available for individuals who will die within the short term compared to those who will not, thereby complicating the accuracy of predictions.

The biomedical and human genetics field has extensively studied model fairness^30–32^, but most studies lack information on sensitive/protected attributes. While electronic health records provide ample information on race and ethnicity, other socioeconomic characteristics are often unavailable, leading to a focus on fairness considering only race/ethnicity, age, and sex in most papers. Our study breaks new ground by comprehensively evaluating fairness across multiple, including multi-level, sensitive/protected attributes. We selected several attributes that are highly valued in Nordic European societies and also are applicable more broadly, including geographical equality, income, marital status, immigration status equality, and destigmatization of mental health diagnoses. For all these attributes we found significantly worse model performance for disadvantaged groups using the RNN model, while none of the differences were significant for the baseline model. Moreover, we observed that being disadvantaged in multiple ways at the same time resulted in substantially worse prediction performance. Several factors, including also those considered as sensitive/protected attributes, are not equally distributed between less densely populated regions compared to more populated regions. For example, previous research suggests that healthcare quality is lower in less densely populated regions^33^, indicating a potential influence on regional disparities. In our study, we observed a positive yet weak association (*r* = 0.23, *p* = 0.05) between population density and AUC in different regional municipalities.

There are different hypotheses proposed to explain why prediction models perform worse for disadvantaged groups across sensitive/protected attributes. One possible explanation is that there are fewer cases in the disadvantaged group, leading to less power during model training^34^. Another explanation is that disadvantaged individuals have lower contact and poorer quality of healthcare, resulting in missing data and measurement errors, ultimately skewing the predictions^11^. Differences in age and sex between socially advantaged and disadvantaged groups could also be an underlying driver of the observed differences in prediction performance, as well as the explicit inclusion of the sensitive/protected attribute as a feature in the model^9^. We thoroughly investigated all these hypotheses by analysing the differences in AUC among monthly pension levels, yet we could not identify the culprit of the variation. We ensured that the number of cases was equal in each bin, and the inclusion of pension information in the model, as well as differences in age and sex distribution between pension bins, did not change the results. While we observed a higher number of socioeconomic records for individuals at higher pension levels, the number of medical records, which we have shown to contribute more to predictive performance, remained comparable across different pension size bins. One possibility is that receiving a higher pension is associated with reduced heterogeneity and entropy. This means that individuals who receive a higher pension may be more similar to each other in terms of contribution of different features to mortality prediction. This also means that cases (i.e., individuals who died within the next year) may stand out more due to the reduced heterogeneity among the controls. This could allow the model to better differentiate between cases and controls, resulting in more accurate predictions.

Our study has several limitations. First, we did not validate the model outside of Finland, highlighting the need for replication in other countries. It would be particularly valuable to assess prediction fairness for socioeconomically disadvantaged groups in different countries, given that Finland has relatively low poverty rates and socioeconomic inequality, as evidenced by a low GINI index, World Bank (2020)^35^. Second, our data lacks biological or genetic markers, self-reported lifestyle information, and other data commonly available in epidemiological studies, but not collected on a nationwide level. The integration of these markers could further improve model performance. Third, most of the fairness analyses were limited to individuals aged 65 and older and to a limited number of sensitive/protected attributes. It is currently unclear what the optimal set of sensitive/protected attributes should be, particularly given the considerable overlap observed in our population. A multidisciplinary approach that includes social scientists and legal experts may be necessary to identify widely available attributes for which AI model fairness should be assessed.

In conclusion, our study demonstrates how deep learning can effectively leverage longitudinal multi-modal nationwide information to accurately predict short-term mortality risk. The model performed well across different causes of death. It also performed well after removing the effect of chronological age, indicating its potential as a population-wide digital marker aging. Future studies should evaluate how probability scores obtained from this model relate to overall health, clinically relevant features and outcomes, as has been done in recent work on a digital marker of coronary artery disease^36^. While there is clear potential for such models, it is important to assess their performance among population groups that already carry the greatest disease burden. We have presented an in-depth examination of fairness at a national scale and revealed that model performance was significantly lower among disadvantaged individuals across multiple sensitive/protected attributes. Therefore, we recommend that studies developing and testing AI models in biomedicine should consider algorithm fairness, entertaining greater integration between socioeconomic and health data.

## METHODS

### Study population

The FinRegistry dataset includes 7,166,416 individuals of whom 5,339,804 (74.51%) are index individuals (every resident in Finland alive on the 1^st^ of January 2010) and the remaining 1,826,612 are relatives (offspring, parents, siblings) and spouses of index individuals, who are not index individuals themselves.

### Inclusion/exclusion criteria

The final sample of this study included alive and not emigrated individuals (*N* = 5,418,753; **Figure 1A**). From an initial sample of 7,166,416, we excluded 1,510,693 individuals who died before predictive intervals of training, validation, and testing sets (**Figure 1B**), 174,948 individuals who emigrated, and 62,022 individuals who have never interacted with healthcare, purchased drugs, or had any entries in socioeconomic registers. These individuals were likely living abroad and given the under-reporting of emigration events (especially within Europe), we excluded these individuals from the study.

### Outcome definition

Our main outcome of interest was mortality. The FinRegistry project has information about individuals’ deaths from two registers Statistics Finland COD and relatives register from Digital and Population Data Services Agency. For our purposes, we considered individuals as deceased if either the year of death was recorded in the SF death register (the year was used because for a small proportion of entries only year, but no exact date was available) or the date of death was recorded in DVV relatives register. Both registers do not fully overlap with larger disagreement in earlier years and considerably smaller in later years. For the period afte^r^ 1st January 2018, there was a good agreement between the two registers (99.83%).

As cases, we considered 54,721 individuals who died during predictive intervals of training, validation, and testing sets (**Figure 1B**). The remaining 5,364,032 were alive during those periods and were considered controls, with a 1.02 case per 100 controls.

### Definition of training, validation, and testing sets

We have randomly split the study population into 3 groups, training (70%), validation (10%), and testing (20%; **Figure 1B**). The first records in the registers used in this study started on th^e^ 1st of January 1969 (the start of the cancer register). Thus, for training purposes, the predictors were considered from th^e^ 1st of January 1969 until a predictive interval which was different for each of the data splits. Validation and testing intervals were shifted one year forward each to allow some external validation in terms of time, leaving validation and testing predi tion periods “unseen” to a model during training This resulted in eature extra tion intervals lasting until 30/09/2017 for training, 30/09/2018 for validation and 30/09/2019 for testing. To increase the model generalizability, we used an external temporal validation approach, where the predictive intervals used to define cases and controls were different for training (1/1/2018-31/12/2018), validation (1/1/2019-31/12/2019) and testing (1/1/2019-31/12/2019). Before each predictive interval, we also left a three-month buffer period, from which no data were not used for training, to avoid potential outcome information leakage into training data.

### Features

Both longitudinal (a) and fixed over time (b) features were considered, with a preference for a longitudinal format which retains more information. Longitudinal features included medical, socio-demographic and geographic records, while fixed over time features included various information predominantly about demographics and health (**Figure 1C**). For a detailed description of features see **Supplementary material**.

### Data preparation and missing data treatment

We kept our data curation to a minimum, for a large part using all medical and sociodemographic records as they appear in original registers, to facilitate transferability and avoid biases which may be introduced with feature engineering. For fixed over time features missing values in continuous and ordinal variables were replaced with mean/mode and an additional binary variable denoting missingness was created. For categorical variables, a category denoting missingness was created. All features were standardized.

### Longitudinal features

For every individual, we considered age as a time scale. That is, all records observed within each year of age were grouped together. The right side of **Figure 1C** shows an illustrative example of how medical and sociodemographic records from each year of an individual’s register history were collated to form sequences used as model inputs. Only unique records within each age year were retained to form a vector of length 100. For a small portion of age year bins (0.03%) that exceeded 100 unique records, a random subsample of 100 values was used and zero-padding was used for the years with fewer than 100 records.

### Fixed over time features

Fixed over time features consisted of categorical, continuous and ordinal features which did not change over time and were not used in a longitudinal fashion within the model. They were instead added separately before the last layer of the model (**Figure 1D**).

### Recurrent neural network model

A good model for sequential health and socioeconomic data should be able to capture complex interactions between records over time. Where the amount of data, sparsity and time windows between records can substantially differ between patients and records could be repeated multiple times. These complexities resemble the challenges also faced in natural language processing (NLP) as individual life events resemble individual words in natural language. Thus, we used a recurrent neural network, namely a gated recurrent unit, which was shown to perform similarly or better than a transformer and other commonly used models with sequential deep learning architecture for clinical event predictions^18,19,37^.

Longitudinally expressed records after embedding were year-by-year of patients’ life used as inputs to a recurrent layer (**Figure 1D**). We follow the TRIPOD recommendations for prediction model development and reporting (see TRIPOD assessment in Supplementary materials).

### Recurrent neural network model‘s hyperparameters

For hyperparameter tuning, we used the Tree-structured Parzen Estimator algorithm implemented within the hyperparameter optimization framework Optuna^38^. For RNN models we have optimised six parameters with the objective to maximise AUC in the validation set. In all the reported analyses, we used the models with an optimised learning rate (lr) of 0.0004, weight decay (L2 penalty) of 7.4*10^−06^, and a dropout rate of 0.46 used in a dropout layer following the RNN layer. The embedding dimension and hidden layer size were 250 and 250. For all models, we used a batch size of 200 as it assured efficient model running given the limited computational resources.

### Baseline model

To evaluate the impact of our deep learning model on performance when compared to only using age and sex information, we used a logistic regression model without any regularization, using only age and sex as features.

### Calibration curves

To assess the calibration of predicted mortality probabilities, we used calibration curves and compared mean predicted probabilities of mortality with observed mortality rates within different predicted probability bins. Ten bins were defined each having an equal number of cases.

### Evaluation of algorithm performance

For binary prediction evaluation, our main metric was the area under the receiver operating characteristic curve (AUC-ROC). This was based on previous literature and clinical recommendations^12,18^. In addition, AUC is not biased towards any class, meaning that both majority and minority classes are equally important when calculating the AUC score. This makes AUC an attractive choice with imbalanced data. However, it is important to note that AUC can still be unreliable when the minority class has an insufficient number of samples. This is because even a small change in the number of correct or incorrect predictions within the minority class can lead to significant changes in the ROC curve and AUC score. To address this issue, we only included sub-samples that had at least 5 samples in the minority class in our analyses. AUC error bars were calculated using bootstrapping. Between-group statistical significance testing was performed using permutation testing by randomly permuting group labels 1000 times.

For survival analyses, we report concordance index and time-dependent AUC at any time between t^he^ 1st t° 52nd week within a predictive interval. We have also split our testing set into three risk groups based on predicted mortality probability: low risk (1-90 percentile), medium risk (91-95 percentile), and high risk (96-100 percentile) and compared the survivability of these groups by plotting Kaplan–Meier curves.

### Fairness evaluation

We chose AUC as our fairness evaluation metric, however, there are many measures that have been suggested to evaluate fairness, with the equalized odds ratio being among the most commonly used^9^. While the equalized odds ratio aims to ensure an equal True Positive Rate (TPR) and False Positive Rate (FPR) between subgroups at a specific probability threshold, AUC parity ensures equal AUCs between subgroups and because ROC curve is a function of FPR and TPR, AUC could be seen as equalized odds ratio at all probability thresholds. Using AUC is especially beneficial for imbalanced samples where choosing a specific probability threshold may be arbitrary. To evaluate fairness, the samples were stratified into subgroups based on their protected attributes. For continuous attributes such as age and pension, we divided subsamples into bins assuring an equal number of cases (individuals who did die during a predictive interval) in each subgroup.

AUC was calculated for each of the stratified subgroups. Additionally, for the pension attribute, we reported accuracy, TPR, and True Negative Rate (TNR). To calculate these measures we used a probability threshold which maximized the geometric mean of sensitivity and specificity: 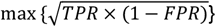.

## Data Availability

Access to FinRegistry data can be obtained by submitting a data permit application for individual-level data for the Finnish social and health data permit authority Findata (https://asiointi.findata.fi/). The application includes information on the purpose of data use; the requested data, including the variables, definitions for the target and control groups, and external datasets to be combined with FinRegistry data; the dates of the data needed; and a data utilization plan. The requests are evaluated on a case-by-case basis. Once approved, the data are sent to a secure computing environment Kapseli and can be accessed within the European Economic Area (EEA) and countries with an adequacy decision from the European Commission.

https://github.com/dsgelab/RNN

https://www.finregistry.fi/finnish-registry-data

## Author contributions

AV and AG wrote the manuscript, with contributions and feedback from all authors. PV and SJ provided valuable clinical perspectives, while SH offered insights from an aging research standpoint. AV and EV preprocessed the FinRegistry data and AL constructed the pedigree for linking relatives in FinRegistry. AV conducted all analyses using the nationwide FinRegistry dataset and produced figures. AG and MP supervised the study.

## Acknowledgements

We are grateful to Finnish individuals, whose data made this study possible. We would also like to thank the entire FinRegistry team for making the data available for the study. Additionally, we would like to acknowledge the researchers who have previously contributed to the development of recurrent neural network models for registry or EHR data, with a special mention to Rasmy et al.^18^, whose openly available code helped our model development process.

This study has received funding from the European Union’s Horizon 2020 research and innovation programme under grant agreement No 101016775, from the European Research Council (ERC) under the European Union’s Horizon 2020 research and innovation program (grant number 945733) and from Academy of Finland fellowship grant N. 323116.

## Data and code availability

Data dictionaries for FinRegistry are publicly available on the FinRegistry website (www.finregistry.fi/finnish-registry-data). Access to FinRegistry data can be obtained by submitting a data permit application for individual-level data for the Finnish social and health data permit authority Findata (https://asiointi.findata.fi/). The application includes information on the purpose of data use; the requested data, including the variables, definitions for the target and control groups, and external datasets to be combined with FinRegistry data; the dates of the data needed; and a data utilization plan. The requests are evaluated on a case-by-case basis. Once approved, the data are sent to a secure computing environment Kapseli and can be accessed within the European Economic Area (EEA) and countries with an adequacy decision from the European Commission.

Essential analysis code used to produce the results is available in the FinRegistry GitHub at: https://github.com/dsgelab/RNN.

