## Supplementary methods and figures for "Deep learning-based prediction of one-year mortality in the entire Finnish population is an accurate but unfair digital marker of aging"

### SUPPLEMENTS

### Supplementary methods

#### Features

Both longitudinal (a) and fixed over time (b) features were considered, with a preference for a longitudinal format which retains more information. Longitudinal features included medical, socioeconomic, and geographic records, while fixed over time features included various information predominantly about demographics and health (Figure 1C).

##### *a. Longitudinal features*

Longitudinal features are represented by medical records (a.1) which can be roughly grouped into predominantly secondary healthcare (a.1.1) and primary healthcare records (a.1.2). Predominantly secondary healthcare data can be classified in curated clinical endpoints (a.1.1.1) and surgical records (a.1.1.2). Primary healthcare records can be classified as ICD-10 records (a.1.2.1), ICPC-2 records (a.1.2.2) and SPAT records (a.1.2.3)

Medication (ATC) records (a.2) capture longitudinal medication purchases. Additional longitudinal data include infectious diseases information (a.3) and longitudinal socioeconomic information (a.4). Socioeconomic information can be categorized into socioeconomic status (a.4.1), occupation (a.4.2), education level and field (a.4.3), and old age pension (a.4.4). Finally, we considered the place of residence (geographic) information (a.5).

##### *a.1 Medical records*

##### *a.1.1 Predominantly secondary healthcare medical records*

Predominantly secondary healthcare medical records were obtained from in-patient and out-patient registers as opposed to primary care registers.

##### *a.1.1.1 Clinical endpoints*

Clinical Endpoints were originally defined for the FinnGen project[1] and later adapted to use within the FinRegistry project. Endpoints were predominantly generated by combining ICD[8-10, O] records coming from Healthcare (HILMO), Causes of death, and Cancer registers. In addition, for a small proportion of endpoints Drug Purchase, Drug Reimbursement, Surgical Procedure, and Primary healthcare ICD records were used. A portion of highly correlated and redundant endpoints was not included, as well as composite endpoints which only included other endpoints but no additional information from clinical registers. Endpoints generated solely from ATC records were also not included as ATC records were used separately. In addition, we have removed endpoints occurring in less than 100 thousand samples. This resulted in 2,860 clinical endpoints used for classification. Clinical endpoints and their definitions can be explored via https://risteys.finregistry.fi/. Clinical endpoints cover the entire study period from the 1st of January 1969.

#### *a.1.1.2 NOMESCO surgical records*

From 1997 surgical procedure records in the HealthCare (HILMO) register were recorded using the NOMESCO classification. Records consist of three alphabetic characters (positions 1-3 of the record) and two numeric characters (positions 4-5 of the records). The first three alphabetic characters denote the functional anatomical body system group, a specific location within a system group, and the method of the procedure. The remaining two numerical characters provide a more fine-grained classification of the procedure. In our study, we have excluded the two last numerical characters as it allowed us to substantially reduce the number of records from 8470 to 2290.

#### *a.1.2 Primary healthcare medical records*

Primary healthcare medical records were obtained from the Primary Healthcare Register (avoHILMO), which started in 2011.

#### *a.1.2.1 ICD-10 records*

ICD-10 records given in the primary care register consist of three to seven hierarchically organised characters. Every record begins with an alphabetical character, which is indicative of the chapter based on a body system. Further two numerical characters broadly define a health condition, and a more refined definition can be given by the remaining characters. In this study, we have used only the first three characters of primary healthcare ICD-10 records and removed rare records which resulted in a total of 1525 records.

#### *a.1.2.2 ICPC-2 records*

ICPC-2 records predominantly encode reasons for primary healthcare visits. The classification is compatible and evolved from ICD classification to better suit primary healthcare needs. It allows recording patients’ reasons for encounters, health problems/diagnoses and primary healthcare procedures and interventions. In this study, we have used a total of 1089 records.

#### *a.1.2.3 SPAT records*

The Finnish classification of functions in outpatient primary healthcare (SPAT) is used to describe functions and procedures in outpatient primary healthcare. In this study, we have used 361 unique 8-symbol SPAT records.

#### *a.2 Medication records*

ATC records classify medicines by active substances in a hierarchical fashion with five different levels. ATC records were obtained from the Drug Purchases register which includes the medicines purchased via pharmacies with a doctor’s prescription and does not include medicines administered during hospital admission. There were 1431 unique ATC records recorded with the register which is available for the 1995-2019 period. We, however, used only the first 5 symbols out of 7 and removed rare ATC records (<1 in 100:000) resulting in 440 unique records retained.

#### a.3 Infectious diseases

The register of infectious diseases is based on disease notifications from medical doctors and laboratories. In our study, we have used information about microbes which caused an infectious disease from 1994 until the predictive interval. In total, there were 170 unique records corresponding to specific microbe groups causing infectious diseases.

#### a.4 Socioeconomic records

In this study, socioeconomic information was used longitudinally as the dynamics of those variables can capture the environmental and social aspects influencing health.

##### a.4.1 Socioeconomic status

Socioeconomic status was based on the information about the main activity, occupation, professional status, and industry of an individual. Eight different socioeconomic status categories (records) were used: lower-level employees, manual workers, upper-level employees, self-employed, students, pensioners, others, and unknown. For individuals for whom the data was available on average socioeconomic status changed 3.4 times. For persons aged 0-15 socioeconomic status was based on the socioeconomic status of the reference person of the household-dwelling. Socioeconomic status was obtained from Statistics Finland and was available from 1970.

##### a.4.2 Occupation

Occupation data was obtained from Statistics Finland and was available from the year 1995 and each occupation was coded hierarchically following the structure of ISCO - International Standard Classification of Occupations[2]. In the register, there were 1277 unique occupations recorded but for our purposes lower level of detail was sufficient and we retained only the first symbol of each record, resulting in 11 unique occupation categories: service and sales workers, professionals, technicians and associate professionals, craft and related trades workers, plant and machine operators, and assemblers, elementary occupations, clerical and support workers, skilled agricultural, forestry and fishery workers, managers, armed forces, unknown.

##### a.4.3 Education

Records for education level and education field were used. Education level is ordered from the lowest to the highest, ranging from secondary to doctoral level education (13 categories in total). The education field based on the International Standard Classification of Education (ISCED) classification contains hierarchical 3-4 symbol records signifying the education area from the broad (first symbol) to the more detailed (further symbols). In total 100 unique education-level records were used. Education information was obtained from Statistics Finland and was available from 1970.

##### a.4.4 Old age pension

Earnings-related old-age pension amounts were taken from the Finnish Centre for Pensions register spanning 1990-2020. The pension amount variable was discretized into a categorical variable with 20 contiguous levels, each having an equal number of samples. It was used longitudinally at each year the pension was received by an individual.

##### *a.5 Place of residence (geographic) information*

Finland has 309 municipalities (2021) and in this study, the geographical location of the individuals was based on the municipality. The geographical information was available from 1964 onwards and to account for changes in municipality definitions (area changes, new municipalities appearing and old disappearing) throughout this follow-up period, the location information was harmonised based on 309 municipalities as defined in 2021. In the models, this information was used longitudinally as individuals for whom the data was available changed the municipality in which they lived on average 2.1 times. As geographic information was available only for index individuals, for individuals born on and after 2010 their mothers' geographic information was used. Living information was obtained from the DVV register.

#### *b. Fixed over time features*

Fixed over time features were grouped into basic demographic and health (b.1), social support (b.2) and birth, relationships, and children (b.3).

##### *b.1. Basic demographic and health*

Basic demographic features predominantly come from DVV registers. However, here we also included basic health features. Number of drug purchases and drug prescriptions from Kela and Kanta registers. Smoking status was recorded in Avohilmo and Birth registers and was available for 30% of the study population (see Supplementary Table 1).

***Supplementary Table 1*: Basic demographic and health fixed over time features.**

| Feature description | Source | Type |
|---|---|---|
| Age in days | DVV | Continuous |
| Sex | DVV Relatives | Binary |
| Index person (living in Finland on 01/01/2010) | DVV | Binary |
| Number of children | DVV Relatives | Ordinal |
| Record(s) in THL Social assistance reg. | THL Soc. Assist. | Binary |
| Record(s) in THL Social Hilmo reg. | THL Soc. Hilmo | Binary |
| Record(s) in THL Infectious diseases reg. | THL Infect. dis. | Binary |
| Record(s) in THL malformations reg. | THL Malformations | Binary |
| Record(s) in THL cancer reg. | THL Cancer | Binary |
| Latest recorded smoking status | THL AvoHilmo + Birth | 6 categories |
| Mother tongue (Fi. / Sw. / Ru. / other / unknown) | DVV Relatives | 5 categories |

##### *b.2. Social support*

We included here information from the THL Register of social assistance about monetary support received by individuals due to lack or insufficiency of income. In addition, here we have included information about the duration of institutional care received by individuals (see Supplementary Table 2).

***Supplementary Table 2*: Social support fixed over time features.**

| Feature description | Source | Type |
|---|---|---|
| Total amount of received social assistance | THL Soc. Assist. | Continuous |
| Duration of social assistance in moths | THL Soc. Assist. | Continuous |
| How many years social assistance spanned | THL Soc. Assist. | Continuous |
| Duration in institutional care | THL Soc. Hilmo | Continuous |

#### *b.3. Birth, relationships, and children*

THL Birth register has a vast amount of information both about newborns and their mothers. Birth information and complications for mothers are recorded in the Hilmo register, however, in that register, there is no such information recorded for newborns. Therefore, here we have included features about a person and their mother at the time of birth and immediately after. Here we also used information from the THL malformation register signifying the severity of malformation and also some features about relationships occurring much later in life (see Supplementary Table 3).

***Supplementary Table 3*: Birth, relationships, and children fixed over time features.**

| Feature description | Source | Type |
|---|---|---|
| Mother's age at birth | THL Birth | Continuous |
| Best estimate of pregnancy duration in days | THL Birth | Continuous |
| Maternal marital status | THL Birth | 9 categories |
| Maternal cohabiting relationship | THL Birth | 3 categories |
| Previous pregnancies | THL Birth | Ordinal |
| Previous miscarriages | THL Birth | Ordinal |
| Previous induced abortions | THL Birth | Ordinal |
| Previous ectopic pregnancies | THL Birth | Ordinal |
| Previous births | THL Birth | Ordinal |
| Previous stillborn births | THL Birth | Ordinal |
| Total number of check-ups | THL Birth | Ordinal |
| Number of outpatient check-ups | THL Birth | Ordinal |
| Mothers smoking status at birth | THL Birth | 5 categories |
| The way in which the child was born | THL Birth | 9 categories |
| Number of foetuses, 1-3 | THL Birth | Ordinal |
| Birth weight, g | THL Birth | Continuous |
| Birth length, cm | THL Birth | Continuous |
| New-born assessment at 7d | THL Birth | 6 categories |
| Score of a Apgar test given to newborns at 1 min | THL Birth | Ordinal |
| Score of a Apgar test given to newborns at 5 min | THL Birth | Ordinal |
| Glucose tested and pathological | THL Birth | Binary |
| Artificial insemination (IVF, ICSI, FET) | THL Birth | Binary |
| Resuscitated immediately after birth | THL Birth | Binary |
| The new-born is resuscitated by the age of 7d | THL Birth | Binary |
| Severity of birth malformation | THL Malformations | 7 categories |
| Ever married | DVV Marriage | 3 categories |
| Ever divorced | DVV Marriage | 3 categories |
| Recorded in birth registry as a mother | THL Birth | Binary |
| Recorded in birth registry as a child | THL Birth | Binary |

### Supplementary Figures

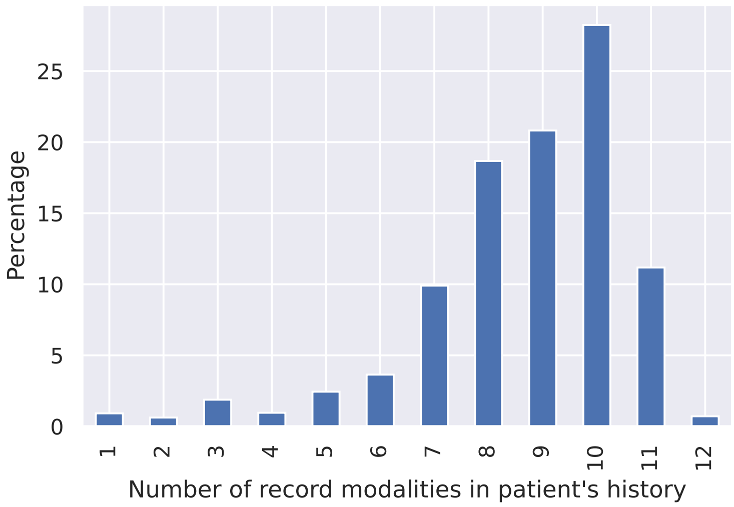

***Supplementary Figure 1*: Distribution of recorded modalities in individual histories.** The chart shows the number of different types of records (modalities) for every individual. Most individuals (78%) had records from 7 to 11 modalities. Education field and level were combined into a single modality since they were always recorded together.

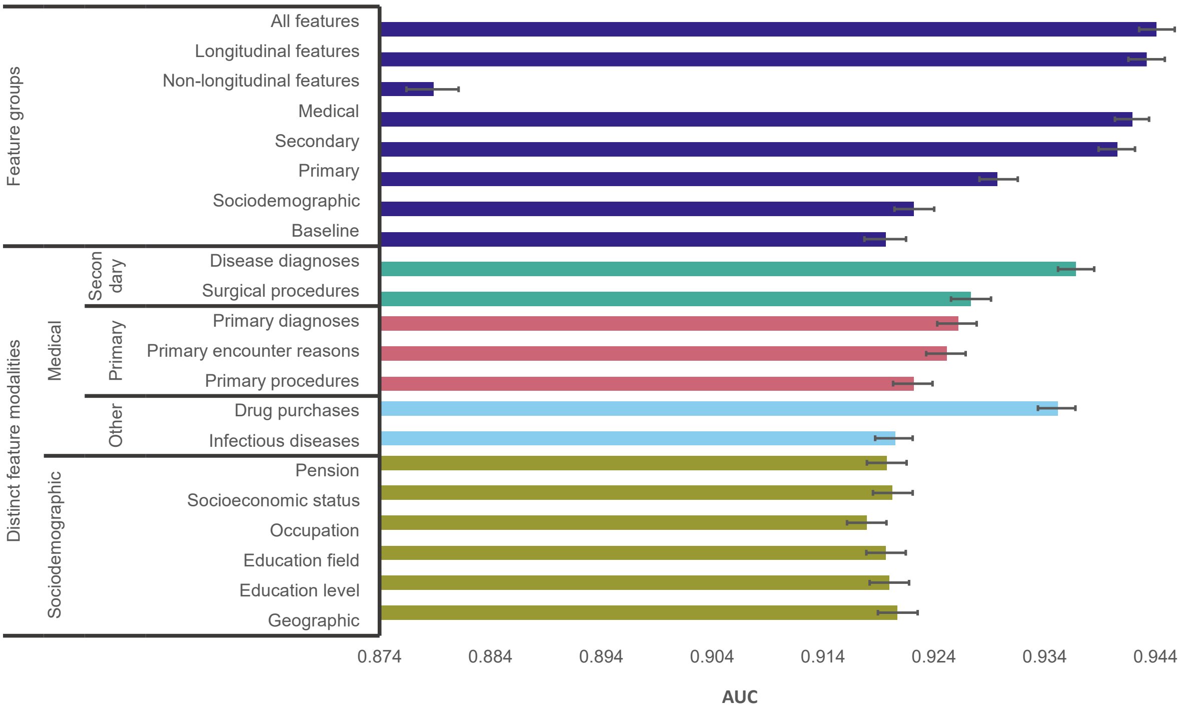

***Supplementary Figure 2*: Importance of feature groups and distinct feature modalities.** Importance was measured by randomly permutating all features except the ones contained within a group or modality of interest. Then the recurrent neural network model was used for classification and areas under the receiver operating characteristic curves were compared. Within feature groups, longitudinal features comprised medical and socioeconomic features.

In turn medical features comprised secondary and primary care features. Non-longitudinal features included fixed over time features. The baseline performance is obtained by permuting all features, which limits the information provided to a model to a certain number of features per age year, while specific feature information for all features is permuted. Classification performance for each individual modality is in the lower part of the figure. The error bars indicate 95% confidence intervals computed using bootstrapping.

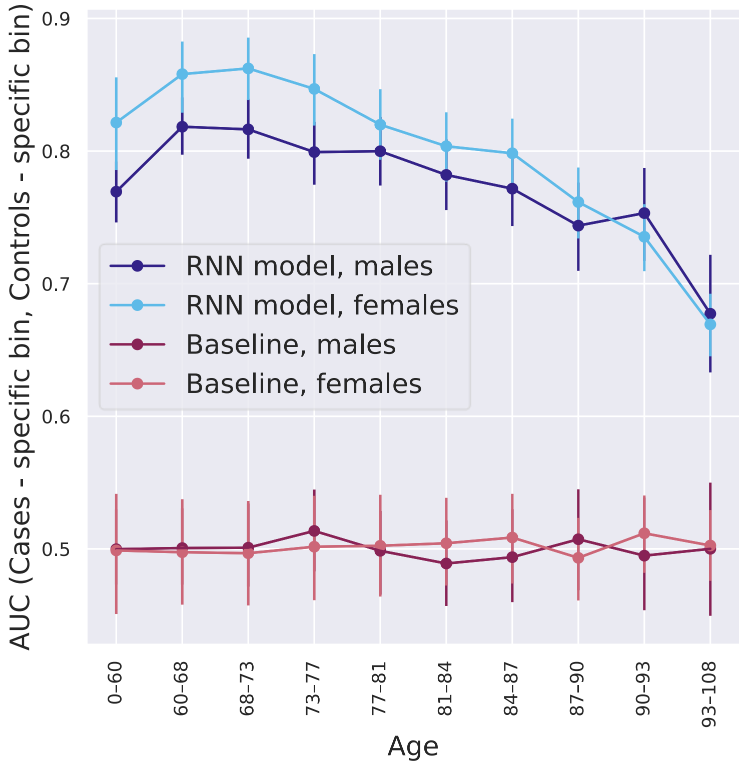

***Supplementary Figure 3*: Areas under the receiver operating characteristic curves within specific age/sex subgroups.**

This figure is comparable to (Figure 4C) and displays areas under the receiver operating characteristic curves within specific age/sex subgroups (with cases and controls from a specific subgroup only). However, here in each bin the controls were matched with respect to age (with a precision level of 1 year). Additionally, the number of controls used was the same as the number of cases. RNN = recurrent neural network.

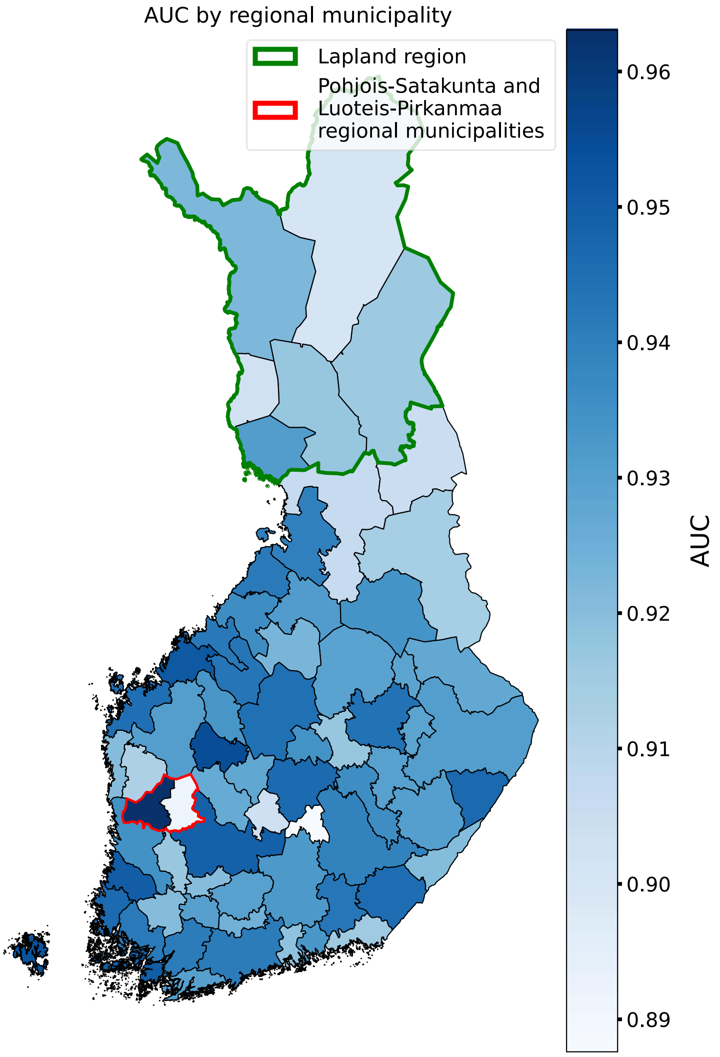

***Supplementary Figure 4*: Variation of area under the receiver operating characteristic curve (AUC) by regional municipality in Finland.**

This figure is like Figure 5A, but the area under the receiver operating characteristic curve (AUC) results presented here are from a model that was trained without using geographic (place of residence) features. The purpose of this was to investigate whether observed geographic differences were due to model's awareness of geographic information. Green border marks Lapland region in which AUC remained significantly lower than in the rest of Finland, $p = 0.004$, Red border surrounds two neighbouring regional municipalities with still significantly different AUCs, $p = 0.002$, Two regional municipalities Mariehamns stad and Ålands skärgård are not plotted because the minority class had less than 20 samples. Statistical significance was assessed using permutation testing.

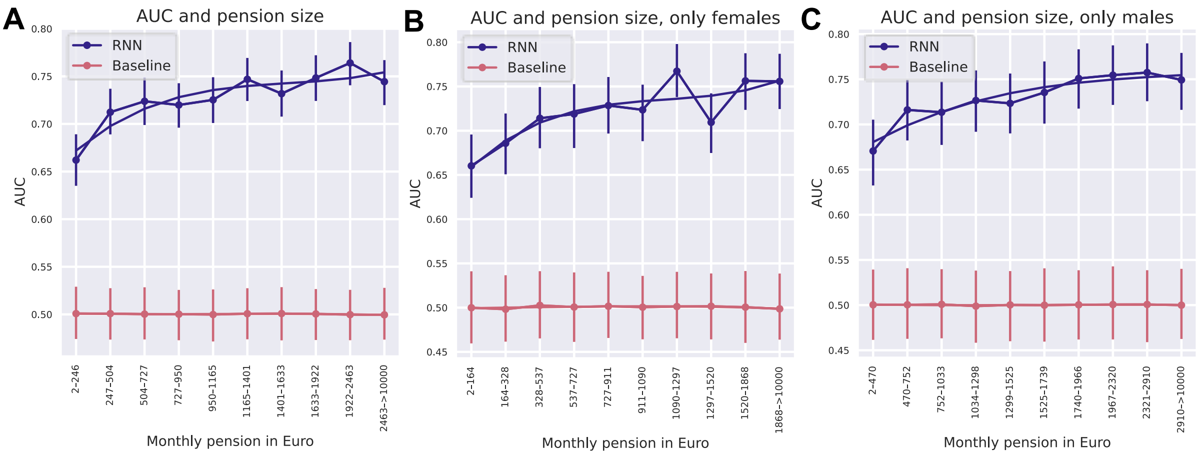

***Supplementary Figure 5*: Areas under the receiver operating characteristic curves (AUC) within specific pension size subgroups.**

(A) This figure is comparable to Figure 5C but here controls were matched for age (with a precision level of 1 year) and sex within each bin. Additionally, the number of controls used was the same as the number of cases. Separate curves were plotted for female (B) and male (C) samples. Despite the significant disparity in pension distribution between sexes, both sexes exhibited comparable AUC differences between different pension bins using the recurrent neural network (RNN) model, as observed in Figure 5C. In contrast, the performance of the baseline model approximated random guessing (≈0.5).

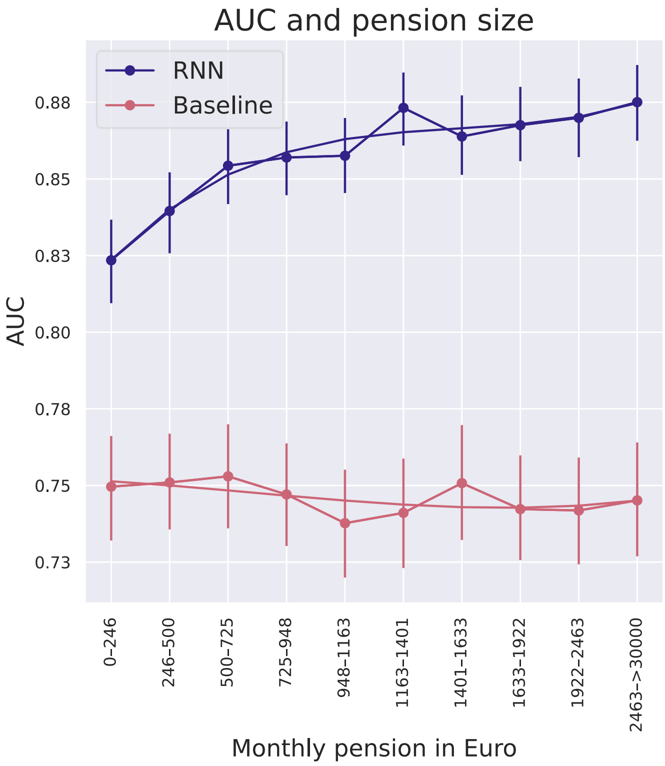

***Supplementary Figure 6*: Areas under the receiver operating characteristic curves (AUC) within specific pension size subgroups without using pension features.**

This figure is like Figure 5C, but the AUC results presented here are from a model that was trained without using pension features. The purpose of this was to investigate whether observed differences in different pension bins were due to model's awareness of pension information. AUCs differences between pension size subgroups remained similar as in Figure 5C. RNN = recurrent neural network.

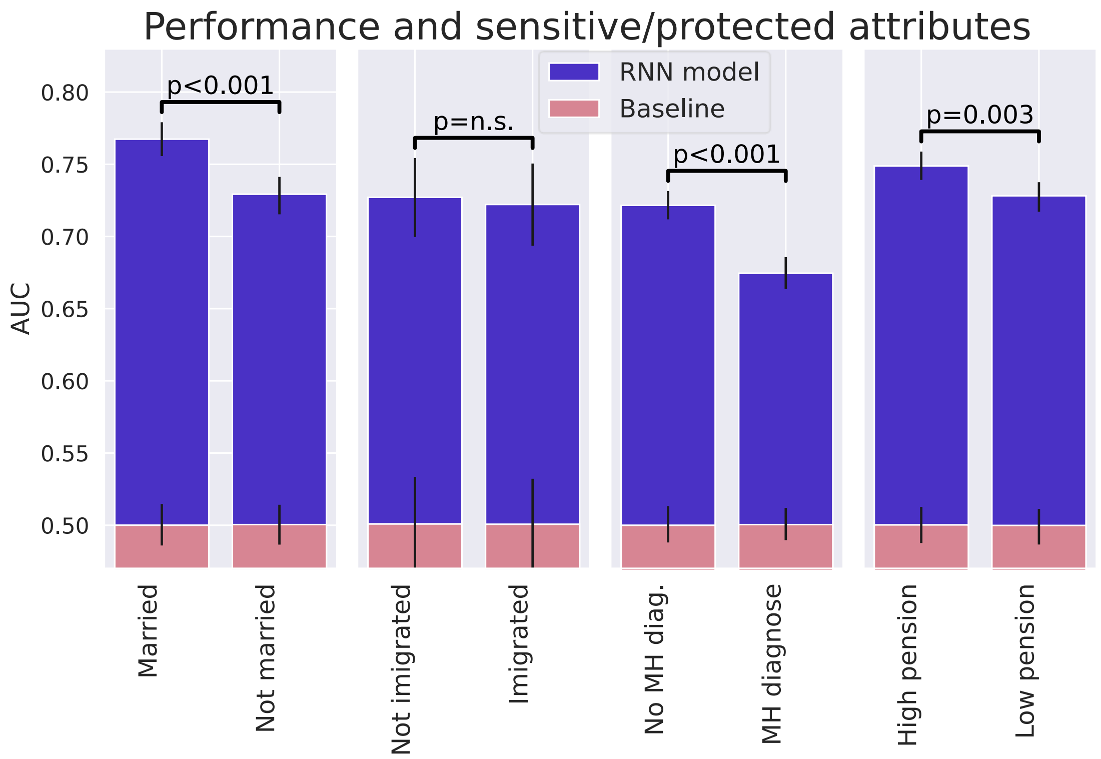

***Supplementary Figure 7*: Areas under the receiver operating characteristic curves (AUC) for different protected or sensitive attributes.**
This figure is comparable to Figure 5F, showing AUCs for different attributes considered protected or sensitive, namely marital status, immigration status, mental health (MH) diagnosis and pension size (individuals were split into two pension size groups assuring an equal number of cases in each). However, in this analysis, socially disadvantaged and advantaged groups were matched for a number of samples, age (with a precision level of 1 year) and sex. Additionally, the number samples, age, and sex of controls were matched to the corresponding cases. Statistical significance was assessed using permutation testing. AUC difference by immigration status became non-significant, likely due to the matching process. This outcome can be attributed to the low number of immigrated individuals who were cases, resulting in limited statistical power. Statistical significance was assessed using permutation testing. RNN = recurrent neural network.

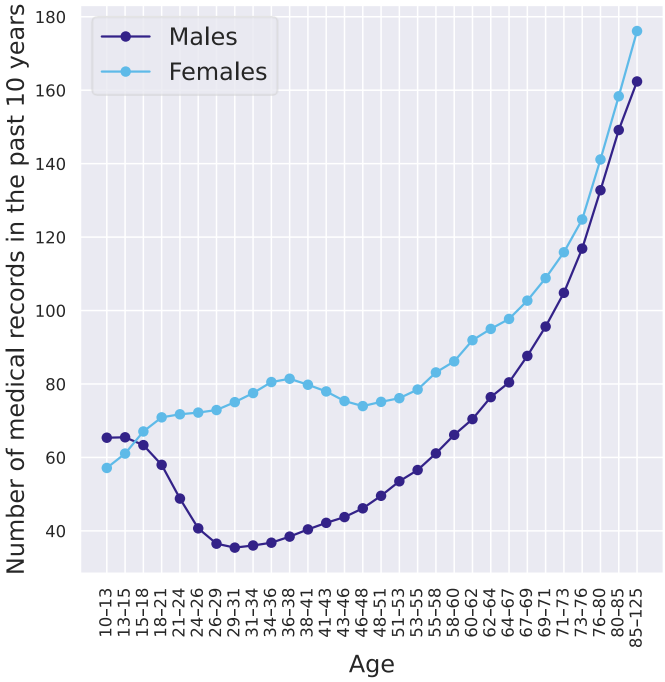

***Supplementary Figure 8*: Amount of data available for different sexes.** Number of medical records recorded from 2010 to 2019 (last 10 years in individuals' medical history), divided by gender, in a test sample.
